# Artificial Intelligence Prediction of Age from Echocardiography as a Marker for Cardiovascular Disease

**DOI:** 10.1101/2025.03.25.25324627

**Authors:** Meenal Rawlani, Hirotaka Ieki, Christina Binder, Victoria Yuan, I-Min Chiu, Ankeet Bhatt, Joseph E. Ebinger, Yuki Sahashi, Andrew P. Ambrosy, Paul Cheng, Alan C. Kwan, Susan Cheng, David Ouyang

## Abstract

Accurate understanding of biological aging and the impact of environmental stressors is crucial for understanding cardiovascular health and identifying patients at risk for adverse outcomes. Chronological age stands as perhaps the most universal risk predictor across virtually all populations and diseases. While chronological age is readily discernible, efforts to distinguish between biologically older versus younger individuals can, in turn, potentially identify individuals with accelerated versus delayed cardiovascular aging. This study presents a deep learning artificial intelligence (AI) approach to predict age from echocardiogram videos, leveraging 2,610,266 videos from 166,508 studies from 90,738 unique patients and using the trained models to identify features of accelerated and delayed aging. Leveraging multi-view echocardiography, our AI age prediction model achieved a mean absolute error (MAE) of 6.76 (6.65 - 6.87) years and a coefficient of determination (R^2^) of 0.732 (0.72 - 0.74). Stratification by age prediction revealed associations with increased risk of coronary artery disease, heart failure, and stroke. The age prediction can also identify heart transplant recipients as a discontinuous prediction of age is seen before and after a heart transplant. Guided back propagation visualizations highlighted the model’s focus on the mitral valve, mitral apparatus, and basal inferior wall as crucial for the assessment of age. These findings underscore the potential of computer vision-based assessment of echocardiography in enhancing cardiovascular risk assessment and understanding biological aging in the heart.

## Introduction

Cardiovascular diseases remain the leading causes of mortality globally, making the accurate assessment of cardiac health essential for effective management and prevention strategies^1^. Echocardiography, a widely used, non-invasive cardiac imaging modality, has played a pivotal role in this effort, offering detailed visualization of cardiac structure and function^2–6^. Advancements in deep learning have opened new avenues for analyzing echocardiographic data, including the prediction of demographic features such as age^7,8^. These predictions, beyond their immediate novelty, carry potential clinical significance by identifying markers of biological aging and indicators of cardiovascular risk.

A key challenge in the training of AI models in medicine is the heterogeneity, imprecision, and lack of labs for model training^9,10^. Most medical imaging has few annotations and focused assessments on narrow diagnoses, even though there is much additional information in medical imaging not identifiable by clinicians^11–13^. Studies are also often constrained by limited datasets, which not only restricts the generalizability of their findings but also acts as a constraint in learning relevant features for an unbiased prediction in deep learning models^14^. In contrast, precise age data is almost always present for medical imaging. While models have largely focused on predicting outcomes like all-cause mortality, such outcomes are limited by few labels, potential loss-of-follow-up, and bias in how datasets are constructed. Less attention has been given to exploring other clinically relevant endpoints or demonstrating the broader predictive capabilities of these systems^15^.

In this study, we address these limitations by developing deep learning models to predict age from multiple echocardiographic views. By using parasternal long axis (PLAX), apical two-chamber (A2C), apical four-chamber (A4C), and subcostal (SC) echocardiographic views for a comprehensive assessment of cardiac form and function, as well as a large well-defined cohort of patients, we sought to evaluate whether AI models can predict age and evaluate whether discordance between predictions and chronological age can be used to understand accelerated and delayed biological aging. We also highlight the potential for the model predictions to predict outcomes beyond all-cause mortality demonstrating the versatility and clinical relevance of this approach.

## Results

### Model Development and Performance

We trained a regression based video ResNet architecture^16^ on each echocardiographic view (PLAX, A2C, A4C, SC) individually. A detailed dataset description is provided in **Table 1**. Evaluation on a held out test set from Cedars-Sinai Medical Center (CSMC) revealed that the individual view models achieved mean absolute errors (MAEs) ranging from 8.62 to 9.85 years (**Table 2)**. These predictions from all four views were then combined using Histogram Gradient Boosting^17^ to create an ensemble model. Notably, the ensemble approach yielded an overall MAE of 6.76 (6.65 - 6.87) years and a coefficient of determination (R^2^) of 0.732 (0.72 - 0.74) on the internal test set from CSMC. While an MAE of 7.200 (7.04 - 7.36) and R^2^ of 0.683 (0.66 - 0.70) was observed for the external test set from Stanford Healthcare (SHC) (**Table 2**).

**Table 1:** Overview of CSMC dataset and SHC datasets.

| Characteristic | CSMC Cohort<sup>1</sup> |  |  | SHC Cohort<br>(External Test)<sup>1</sup> |
| --- | --- | --- | --- | --- |
|  | Training | Validation | Test |  |
| **Patients** | 85,755 | 897 | 4,086 | 4,786 |
| **Studies** | 155,243 | 1,685 | 9,580 | 5,528 |
| **Videos** | 2,436,040 | 26,239 | 147,987 | 76,632 |
| **PLAX** | 873,198 (35.81%) | 9,479 (36.08%) | 53,633 (36.20%) | 27,114 (35.38%) |
| **A4C** | 834,047 (34.20%) | 8,889 (33.84%) | 50,512 (34.09%) | 28,432 (37.10%) |
| **SC** | 401,154 (16.45%) | 4,392 (16.72%) | 23,888 (16.12%) | 13,218 (17.25%) |
| **A2C** | 329,998 (13.53%) | 3,506 (13.34%) | 20,135 (13.60%) | 7,868 (10.27%) |
| **Age (years)** | 65.97 ± 17.23 | 65.46 ± 16.42 | 66.78 ± 16.96 | 60.96 ± 16.69 |
| **Female Count (n, %)** | 1,155,284 (47.38%) | 12,221 (46.53%) | 62,416 (42.13%) | 34,447 (44.95%) |
| **Race** |  |  |  |  |
| **White (n, %)** | 1,651,213 (67.78%) | 17,392 (66.33%) | 101,974 (68.84%) | 41,551 (54.22%) |
| **Asian (n, %)** | 174,632 (7.17%) | 1,928 (7.36%) | 9,838 (6.65%) | 12,527 (16.34%) |
| **Black or African American (n, %)** | 379,596 (15.59%) | 4,268 (16.28%) | 22,170 (14.98%) | 4,284 (5.59%) |
| **Other (n, %)** | 232,956 (9.56%) | 2,678 (10.21%) | 14,186 (9.59%) | 18,270 (23.84%) |
| **LVEF < 41 (n, %)** | 366,729 (15.04%) | 3,313 (12.61%) | 23,781 (16.05%) | 9,410 (12.28%) |
| **LVEF (%)** | 56.79 ± 15.03 | 58.07 ± 13.94 | 56.10 ± 15.41 | 54.33 ± 12.92 |
| **Aortic Regurgitation (n, %)** | 507,170 (20.80%) | 4,797 (18.26%) | 31,415 (21.20%) | 14,657 (19.13%) |
| **Mitral Regurgitation (n, %)** | 1,019,600 (41.81%) | 10,607 (40.38%) | 65,408 (44.14%) | 31,734 (41.41%) |
| **Tricuspid Regurgitation (n, %)** | 1,149,778 (47.15%) | 11,716 (44.61%) | 74,395 (50.21%) | 37,806 (49.33%) |
| **Aortic Stenosis (n, %)** | 169,753 (6.96%) | 1,570 (5.98%) | 12,883 (8.69%) | 3,899 (5.10%) |
| **BMI** | 27.25 ± 8.85 | 27.65 ± 8.01 | 27.10 ± 7.18 | 26.94 ± 6.42 |
| **LVIDd 2D (cm)** | 4.53 ± 1.82 | 4.50 ± 1.16 | 4.55 ± 1.08 | 4.75 ± 0.86 |
| **LVIDs 2D (cm)** | 3.14 ± 1.06 | 3.09 ± 0.88 | 3.18 ± 1.06 | 3.34 ± 0.96 |
| **LVPWd 2D (cm)** | 1.08 ± 1.20 | 1.09 ± 0.29 | 1.09 ± 0.31 | 1 ± 0.21 |
| **IVSd 2D (cm)** | 1.10 ± 0.54 | 1.10 ± 0.30 | 1.13 ± 0.34 | 1.03 ± 0.24 |
| **TR Peak Gradient (mmHg)** | 26.75 ± 12.91 | 24.91 ± 11.08 | 27.23 ± 12.66 | 29.54 ± 13.46 |

<sup>1</sup> (mean ± std.dev.); (n, %). BMI: Body Mass Index, LVIDd 2D: Left Ventricular Internal Dimension in Diastole, 2D, LVIDs 2D: Left Ventricular Internal Dimension in Systole, 2D, LVPWd 2D: Left Ventricular Posterior Wall Dimension in Diastole, 2D, IVSd 2D: Interventricular Septum Dimension in Diastole, 2D, TR Peak Gradient: Tricuspid Regurgitation Peak Gradient.

**Table 2:** Performance metrics for each echocardiographic view evaluated on the CSMC test split and the external SHC cohort. The model was trained on the CSMC cohort, excluding patients who had undergone major cardiac procedures (CABG, heart transplant, or valve replacement)

| Echocardiography View | CSMC Cohort |  | SHC Cohort |  |
| --- | --- | --- | --- | --- |
|  | MAE | R<sup>2</sup> | MAE | R<sup>2</sup> |
| **PLAX** | 8.662 (8.6 - 8.72) | 0.562 (0.55 - 0.59) | 8.342 (7.72 - 8.14) | 0.575 (0.57 - 0.58) |
| **A4C** | 8.62 (8.56 - 8.68) | 0.569 (0.56 - 0.58) | 9.46 (9.36 - 9.56) | 0.434 (0.42 - 0.45) |
| **A2C** | 9.016 (8.91 - 9.12) | 0.521 (0.51 - 0.53) | 8.479 (8.32 - 8.64) | 0.544 (0.53 - 0.56) |
| **SC** | 9.851 (9.75 - 9.94) | 0.456 (0.44 - 0.46) | 10.399 (10.26 - 10.54) | 0.351 (0.34 - 0.37) |
| **Ensemble** | 6.76 (6.65 - 6.87) | 0.732 (0.72 - 0.74) | 7.200 (7.04 - 7.36) | 0.683 (0.66 - 0.70) |

### Survival Analysis Based on Predicted Age Groups

We stratified patients based on their predicted age to explore associations with clinical outcomes. Kaplan-Meier survival curves^18^ for 5-year follow-up period revealed a clear separation between groups, with studies predicted to be in the older age groups showing significantly worse survival. This separation was also pronounced for coronary artery disease, heart failure, and stroke (**Figure 1)**^19^. Statistical analysis confirmed that these differences were highly significant across all outcomes (p < 0.001). To compare the predictive ability of chronological and biological age, we also performed stratification using chronological age (**Supplementary Figure 1**).

**Figure 1:**
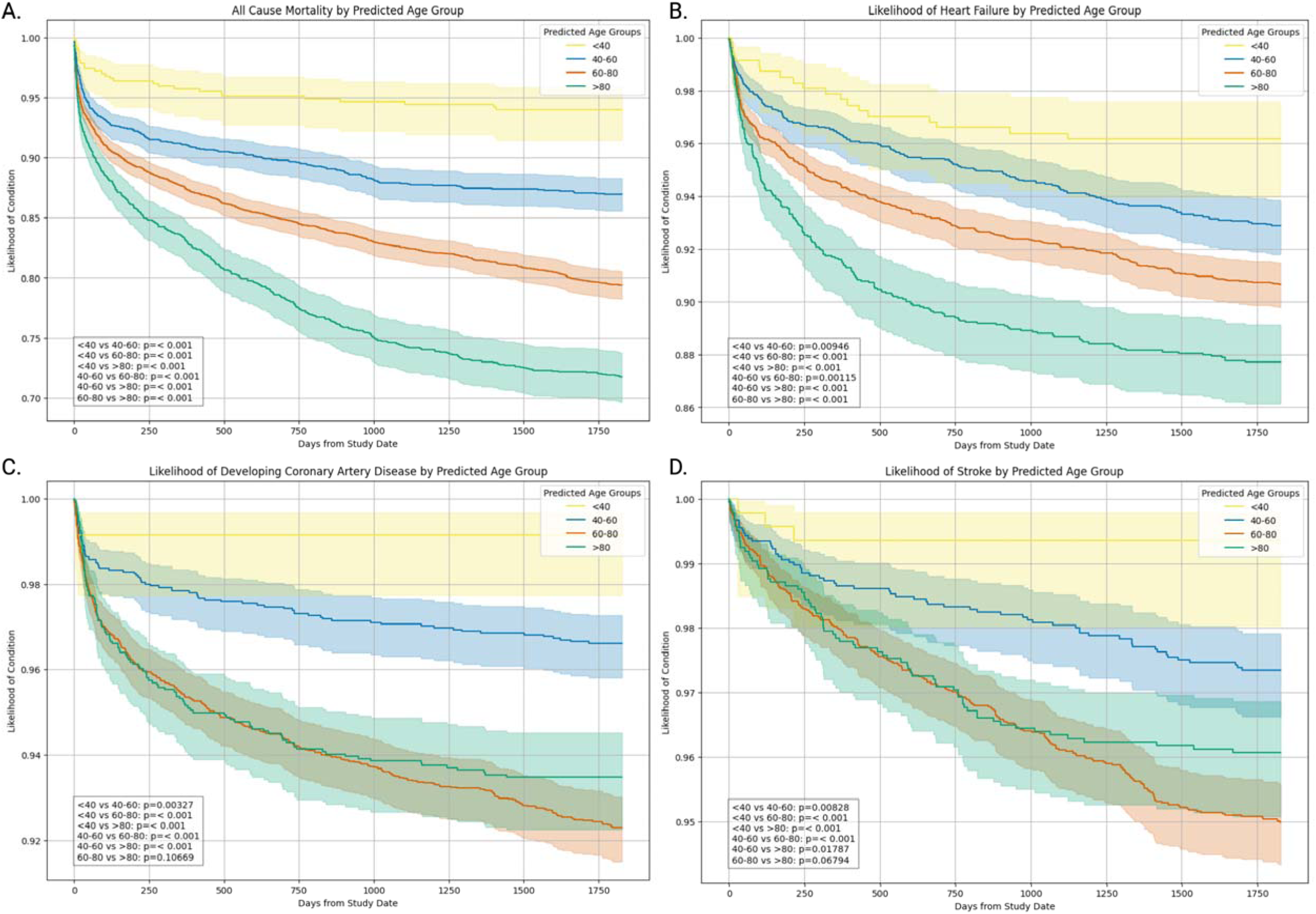
Kaplan-Meier survival curves illustrating the association between predicted age group and four clinical endpoints: (A) all-cause mortality, (B) heart failure, (C) coronary artery disease, and (D) stroke. Each panel displays the survival probability over 5 years for groups stratified by predicted age, with steeper declines indicating higher event rates.

We also analyzed the association between age, predicted age, and key cardiovascular outcomes, adjusting for relevant clinical predictors including smoking status, diabetes mellitus, systolic blood pressure, high-density lipoprotein cholesterol, total cholesterol, left ventricular ejection fraction (LVEF), valvular heart disease and prior cardiovascular interventions. Hazard ratios (HRs) were computed per 10-year increase in both chronological age and predicted age (**Figure 2B**). For all-cause mortality, while chronological age demonstrated an association (HR = 1.33, 95% CI: 1.28–1.37, p < 0.005), predicted age exhibited a stronger effect (HR = 1.42, 95% CI: 1.37–1.48, p < 0.005). Similar trends were observed for coronary artery disease (CAD) (chronological age: HR = 1.15, 95% CI: 1.09–1.21, p < 0.005; predicted age: HR = 1.25, 95% CI: 1.17–1.34, p < 0.005), heart failure (chronological age: HR = 1.16, 95% CI: 1.10–1.21; predicted age: HR = 1.21, 95% CI: 1.14–1.28, p < 0.005), and stroke (chronological age: HR = 1.07, 95% CI: 1.01–1.15, p = 0.03; predicted age: HR = 1.16, 95% CI: 1.06–1.26, p < 0.005). Across all outcomes, predicted age demonstrated a higher HR compared to chronological age.

**Figure 2:**
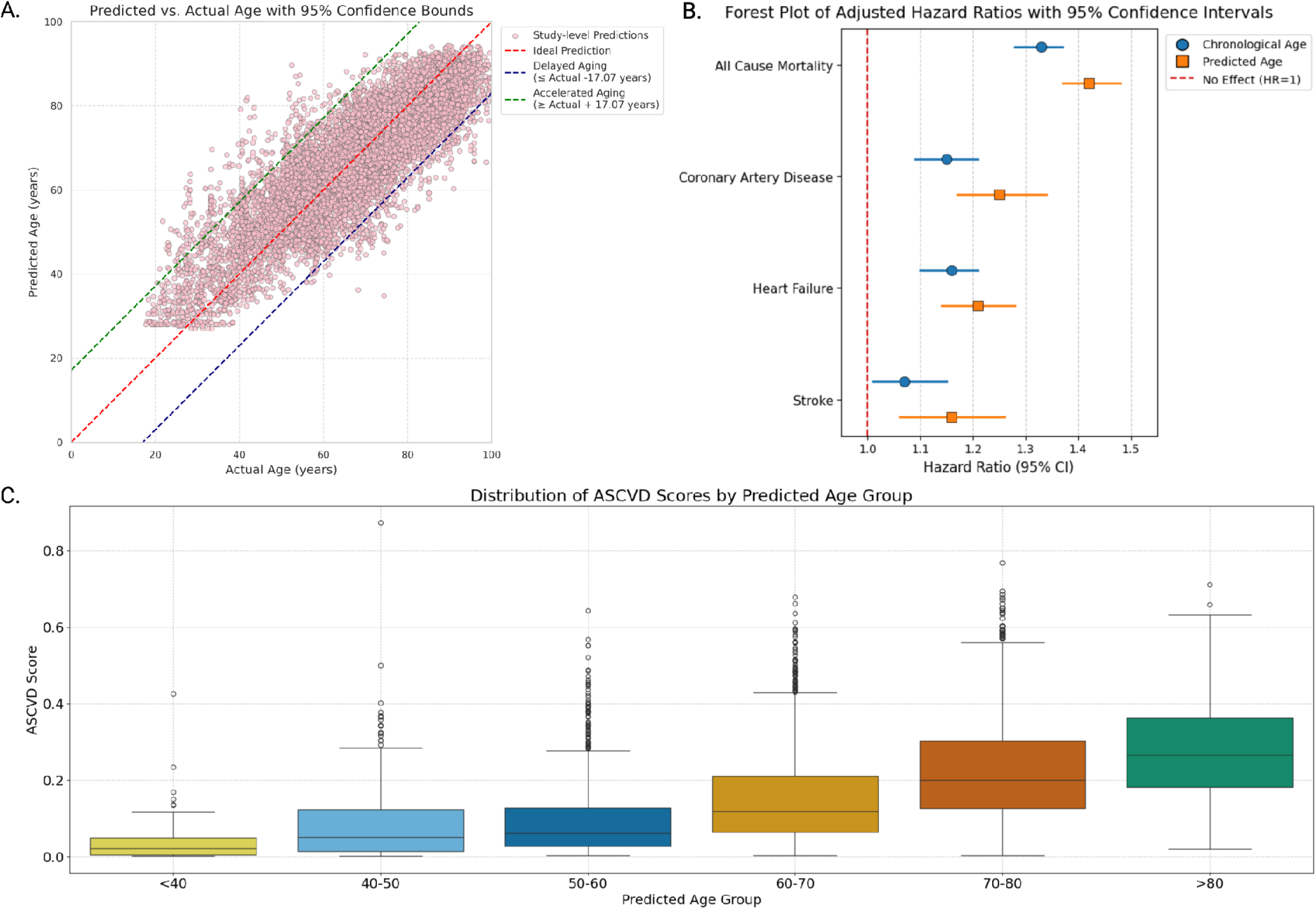
**(A)** Scatter plot of predicted vs. chronological age. The red diagonal line indicates normal aging; dashed lines show accelerated (above) and delayed (below) aging. Each dot represents a study. **(B)** Forest plot of adjusted hazard ratios (HRs) with 95% confidence intervals for age and predicted age across cardiovascular outcomes. **(C)** Box plots of ASCVD risk scores by six predicted age groups. Boxes show IQRs with medians; whiskers represent 1.5×IQR, and outliers are shown as individual points.

### Predicted Cardiac Aging Phenotypes and Their Association with Valvular Conditions

Based on the ensemble model predictions, we classified our study cohort into three cardiac aging phenotypes: “Accelerated”**, “**Expected”, and “Delayed” based on the difference between chronological age and predicted age (**Figure 2A**). Accelerated cardiac aging was defined as having a residual (Predicted Age – Chronological Age) below the lower 95% confidence interval limit, where the predicted age was significantly higher than the chronological age. Delayed cardiac aging was defined as having a residual above the upper limit, where the predicted age was much lower than the chronological age. Expected cardiac aging was characterized by residuals within the confidence interval, where the predicted age closely matched the chronological age.

We found a clear trend of increasing prevalence of valvular heart disease as the cohort’s predicted age group progresses (**Table 3**). Specifically, individuals classified as having accelerated cardiac aging exhibited higher rates of aortic stenosis as well as aortic, mitral and tricuspid regurgitation compared to those in the expected and delayed aging groups. Conversely, individuals in the delayed aging category generally showed significantly lower rates of these conditions across most predicted age groups.

**Table 3:** Chi-square test results for the association between predicted and chronological age groups across various valvular abnormalities (stenosis and regurgitation) in different predicted age categories.

| Predicted Age Group | Valvular Disorders | Predicted age significantly lower than chronological age | Prediction within expected range | Predicted age significantly higher than chronological age | Chi<sup>2</sup> Statistic | Degrees of Freedom | p-value |
| --- | --- | --- | --- | --- | --- | --- | --- |
| < 40 | Aortic Stenosis | - | 3 (0.50%) | 5 (3.97%) | 8.59 | 1 | 0.003 |
|  | Mitral Regurgitation | - | 96 (15.92%) | 45 (35.71%) | 24.92 | 1 | <0.001 |
|  | Tricuspid Regurgitation | - | 185 (30.68%) | 62 (49.21%) | 15.15 | 1 | <0.001 |
|  | Aortic Regurgitation | - | 37 (6.14%) | 26 (20.63%) | 25.94 | 1 | <0.001 |
| 40 - 60 | Aortic Stenosis | 0 (0.00%) | 40 (1.84%) | 17 (11.26%) | 52.75 | 2 | <0.001 |
|  | Mitral Regurgitation | 1 (12.50%) | 615 (28.29%) | 67 (44.37%) | 18.73 | 2 | <0.001 |
|  | Tricuspid Regurgitation | 3 (37.50%) | 799 (36.75%) | 88 (58.28%) | 27.73 | 2 | <0.001 |
|  | Aortic Regurgitation | 0 (0.00%) | 220 (10.12%) | 43 (28.48%) | 48.59 | 2 | <0.001 |
| 60 - 80 | Aortic Stenosis | 0 (0.00%) | 260 (6.40%) | 3 (15.00%) | 9.37 | 2 | 0.009 |
|  | Mitral Regurgitation | 7 (7.00%) | 1590 (39.15%) | 12 (60.00%) | 46.55 | 2 | <0.001 |
|  | Tricuspid Regurgitation | 31 (31.00%) | 1882 (46.34%) | 14 (70.00%) | 13.87 | 2 | <0.001 |
|  | Aortic Regurgitation | 3 (3.00%) | 721 (17.75%) | 9 (45.00%) | 25.18 | 2 | <0.001 |
| > 80 | Aortic Stenosis | 14 (13.59%) | 451 (20.58%) | - | 2.56 | 1 | 0.11 |
|  | Mitral Regurgitation | 46 (44.66%) | 1303 (59.47%) | - | 8.31 | 1 | 0.004 |
|  | Tricuspid Regurgitation | 63 (61.17%) | 1507 (68.78%) | - | 2.3 | 1 | 0.129 |
|  | Aortic Regurgitation | 30 (29.13%) | 810 (36.97%) | - | 2.28 | 1 | 0.131 |

### Age Prediction in Heart Transplant Patients

We further investigated age-related patterns in heart transplant patients by analyzing the relationship between chronological age and model predicted age. We calculated the mean age of heart transplant patients (n = 53) within 1 year before and after the transplant. For the pre-transplant period, the mean age was 62.57 ± 7.57 years, while for the post-transplant period, the mean age decreased to 57.32 ± 7.78 years (t-statistic = 4.81, p-value < 0.001), indicating a substantial reduction in mean predicted age following the transplant. Importantly, 37 of the 50 patients who had studies both before and after the transplant in the one-year period were predicted to be younger following the transplant. Our analysis revealed that the model frequently predicted patients to be younger following heart transplantation, a trend reflected in **Supplementary Figure 2**.

### Atherosclerotic Cardiovascular Disease Risk Score Across Predicted Age Groups

To further evaluate the clinical relevance of our model’s age predictions, we grouped studies according to their predicted age (<40, 40–50, 50–60, 60–70, 70–80 and >80) and compared their atherosclerotic cardiovascular disease (ASCVD) risk scores^20^ (**Figure 2C**). Out of the 9,580 studies in the test set, we were able to calculate the ASCVD score for 4,773 studies, as traditional ASCVD risk scores typically exclude individuals under 40, over 80, and those with pre-existing ASCVD. Additionally, certain parameters were unavailable for some studies, further limiting score calculation. We observed an incremental rise in mean 10-year ASCVD risk with each predicted age group. For individuals predicted to be under 40 years (54 studies), the mean 10- year ASCVD risk was 4.6%. In the predicted 40–50 age group (427 studies), the mean risk increased to 8.3%. The predicted 50–60 age group (1,140 studies) had a further increase to 9.6%, while those predicted 60–70 years (1,726 studies) had a mean 10-year ASCVD risk of 15.2%. In the predicted 70–80 age group (1,154 studies), the mean risk rose significantly to 22.6%. Finally, for individuals predicted to be over 80 (272 studies), the mean 10-year ASCVD risk was 27.8%.

### Interpretability

To evaluate the most relevant cardiac structures, we employed guided backpropagation^21^ to visualize regions of interest within each echocardiographic view that contributed most significantly to the model’s predictions (**Figure 3**). The saliency maps indicated that the model concentrated predominantly on the regions of the crux cordis, mitral valve, and the mitral annulus across multiple views, suggesting consistent regions of interest that highlight important cardiac structures.

**Figure 3:**
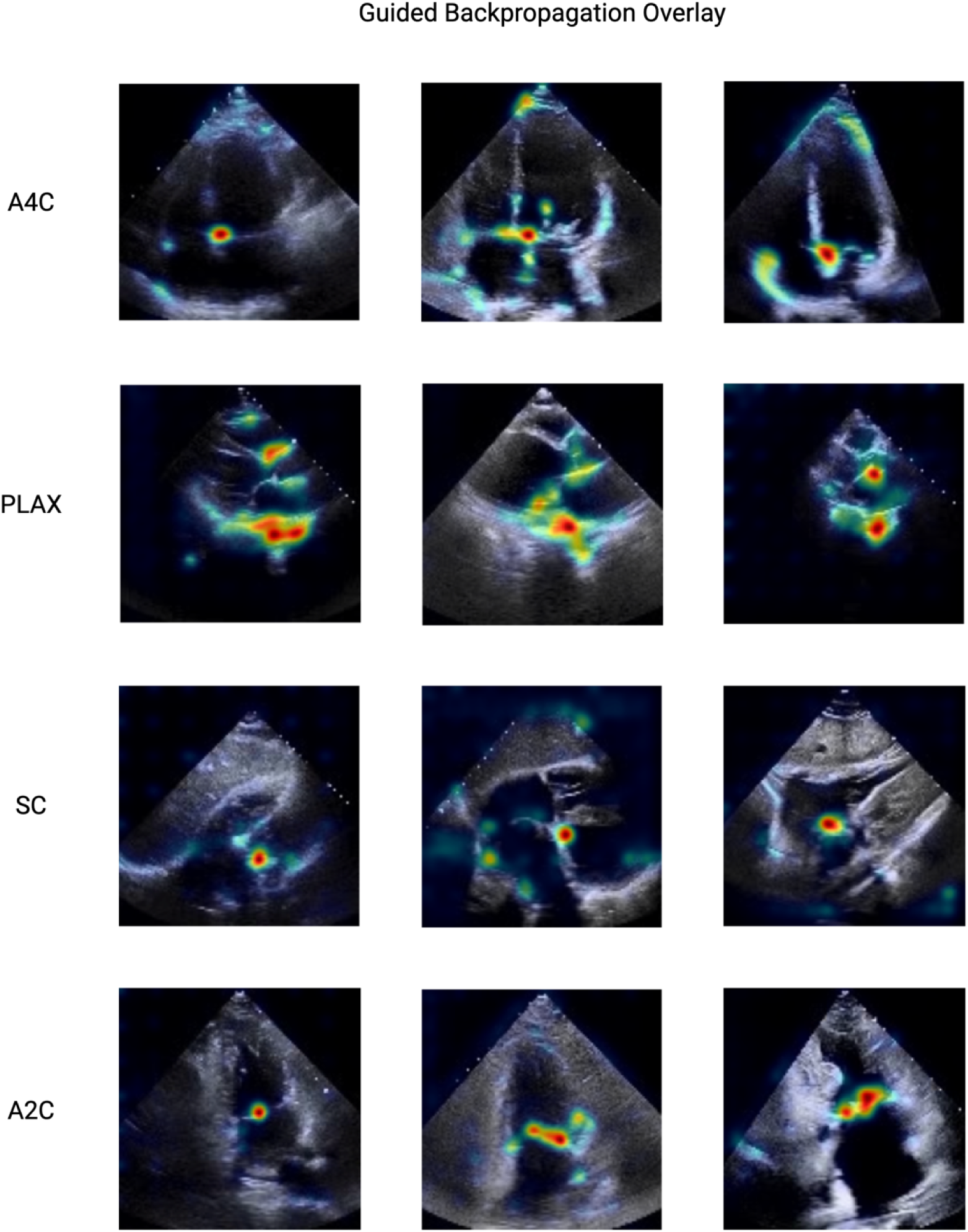
Guided backpropagation interpretability method highlighting the most important regions of interest for the model to make determination of age.

## Discussion

Our study demonstrates that deep learning models can accurately predict age from echocardiogram videos, potentially serving as a proxy for biological cardiac aging. A key aspect of our analysis is the interpretation of residuals and discrepancies between predicted and chronological age, as ability to identify individuals whose predicted age deviates from their chronological age could offer valuable insights into cardiovascular health. Furthermore, by comparing saliency maps across multiple cardiac views and images showing different anatomic structures, we identify key cardiac structures that change over time and with accelerated aging. Our findings suggest that these imaging features of aging may capture clinically significant aging processes.

The distinct separation observed in Kaplan-Meier survival curves for overall survival, coronary artery disease, heart failure, and stroke when classifying patients into accelerated or delayed aging underscores the clinical relevance of our model’s predictions. This provides strong evidence that predicted age serves as a valuable prognostic tool for cardiovascular risk, outperforming stratification by chronological age as an alternative. Across all outcomes, predicted age consistently demonstrated a higher hazard ratio, suggesting that predicted age estimates capture additional risk information beyond traditional age-based assessments. Additionally, the incremental rise in mean ASCVD risk scores across predicted age groups further validates our model’s clinical relevance^20^ and underscores the potential of integrating our model for more accurate risk stratification and prognosis.

An intriguing aspect of our findings is the "rejuvenation" effect observed in post-heart transplant patients, where the model predicted younger ages following transplant. This observation aligns with the model’s focus on age-sensitive cardiac features, suggesting that post-transplant cardiac morphology and function resemble that of a younger heart, despite the recipient’s chronological age ^25^.

The guided backpropagation saliency maps revealed that our model focuses on clinically relevant cardiac structures, primarily the crux cordis, mitral annulus, and mitral valve across all views. The mitral annulus is a prominent region where age-related structural and functional changes, such as mitral annular calcification, commonly occur^26^. The mitral valve is also prone to degenerative changes like sclerosis or calcification with age ^26,27^. This alignment lends biological plausibility to our model’s predictions and enhances its interpretability for clinicians.

Despite these promising findings, several limitations warrant consideration. First, while the dataset was extensive, it originated from a single institution, which may limit generalizability. Future studies should validate these models across diverse populations and institutions^29^. Second, although saliency maps provided valuable insights into model focus areas, more sophisticated explainability techniques could further elucidate the specific features driving predictions^30^. Also, the echocardiographic videos analyzed are likely from patients presenting with health concerns, as they sought hospital care, potentially introducing a bias that may not represent normal aging, even after excluding patients with major procedures.

In conclusion, this study highlights the potential of deep learning models applied to echocardiographic data for predicting biological age and assessing cardiovascular risk. By providing interpretable insights and demonstrating clinical relevance, this approach paves the way for more personalized and proactive cardiovascular care.

## Methods

### Cohort Selection

Echocardiography studies spanning from 2012 to 2022 were obtained from the CSMC. The videos were originally acquired using GE or Philips ultrasound machines, stored as DICOM files, and converted into AVI format^32^. These videos files underwent pre-processing to ensure consistency and extraneous details beyond the ultrasound section were removed, metadata was de-identified, and the videos were resized to resolution of 112 × 112 to facilitate computational efficiency.

The dataset included videos from four standard echocardiographic views: A4C, A2C, PLAX, and SC. Doppler videos were excluded from the dataset. Each video was filtered to include at least 32 frames, from randomly sampled 32 frames, every other frame (16 frames per clip) was inputted into the model. The studies also underwent additional filtering to exclude patients with prior histories of coronary artery bypass grafting (CABG), cardiac valve replacement, or heart transplant from the training and the validation splits. The demographics, diagnoses, and procedural history was extracted from the electronic health record, where it is stored by international classification of disease (ICD) codes **(Supplementary Table 2)**. This exclusion was done to ensure model learned features without confounding effects from such interventions. The dataset was divided into training, validation, and test splits based on unique patients to prevent data leakage. A detailed overview of the included studies is presented in **Table 1**. Separately, a cohort of 76,632 videos from 5,528 studies of 4,786 unique patients from SHC were used as an external validation set (**Table 1**). This study was approved by the Institutional Review Boards at CSMC and SHC.

### Model Description and Implementation Details

The spatiotemporal relationships within echocardiographic videos were captured using a deep learning model based on the ResNet architecture (R2 + 1D)^16^. This model processes 3D video data using a combination of 2D spatial convolutions and 1D temporal convolutions at every convolutional layer. The architecture was chosen to balance computational efficiency with the ability to capture dynamic cardiac features across both space and time. Models were trained using stochastic gradient descent with the ADAM optimizer, with an initial learning rate of 0.01, momentum of 0.9, and a learning rate decay schedule.

Deep learning models were trained separately for each echocardiographic view (A4C, A2C, PLAX, and SC). To further improve performance, an ensemble model was constructed using a gradient boosting regressor^17^. The inputs to the ensemble model were predictions from the individual view-specific models, generated by running inference on the validation set. This allowed the ensemble model to leverage complementary information from different echocardiographic views while making sure that the same test set was used for evaluating both individual and ensemble models, ensuring a fair comparison of performance.

### Statistical analysis

Model performance was assessed through mean absolute error (MAE) and the coefficient of determination (R²) for the age prediction task, using both the test and external validation datasets. Stratified analyses were conducted based on predicted age groups to explore clinical outcomes. The Z-score of 1.96 is used to calculate the 95% confidence interval (CI) by multiplying it with the standard deviation of the residuals (chronological age – predicted age). The upper bound (accelerated aging) is the mean residual plus this CI, and the lower bound (delayed aging) is the mean residual minus this CI. Kaplan-Meier survival curves were employed to assess the separation between groups for overall survival, coronary artery disease, heart failure, and stroke. The significance of these differences was evaluated using log-rank tests for pairwise comparisons and a multivariate log-rank test to assess overall differences across all age groups. A multivariate Cox proportional hazards model to assess the association between chronological age and predicted age and the time-to-event outcome. Covariates included smoking status, diabetes mellitus, systolic blood pressure, high-density lipoprotein cholesterol (hdlc), total cholesterol, left ventricular ejection fraction (LVEF), valvular heart disease and prior cardiovascular interventions and were selected based on clinical relevance. Hazard ratios (HR) and 95% confidence intervals (CI) were reported to quantify the effects of each variable on survival outcomes. Pooled Cohort Equations were used to calculate the ASCVD risk score. Statistical analysis was performed with Python (version 3.10.12), and 95% confidence intervals were computed based on 10,000 bootstrap samples.

## Data Availability

Data available on reasonable request

## Code and Data Availability

The code and model weights are available at https://github.com/echonet/aging. The patient data is not publicly available due to their potentially identifiable nature.

## Author contributions

Concept and design: MR, DO Code and programming: MR, HI

Data acquisition, analysis, or interpretation of data: MR, IC, VY, YS, CB, DO

Drafting and Critical revision of the manuscript: MR, CB, AB, SC, APA, ACK, JEE, PC, SC, DO

Statistical analysis: MR, HI, DO Obtained funding: DO Supervision: DO

---

**Supplementary Figure 1:**
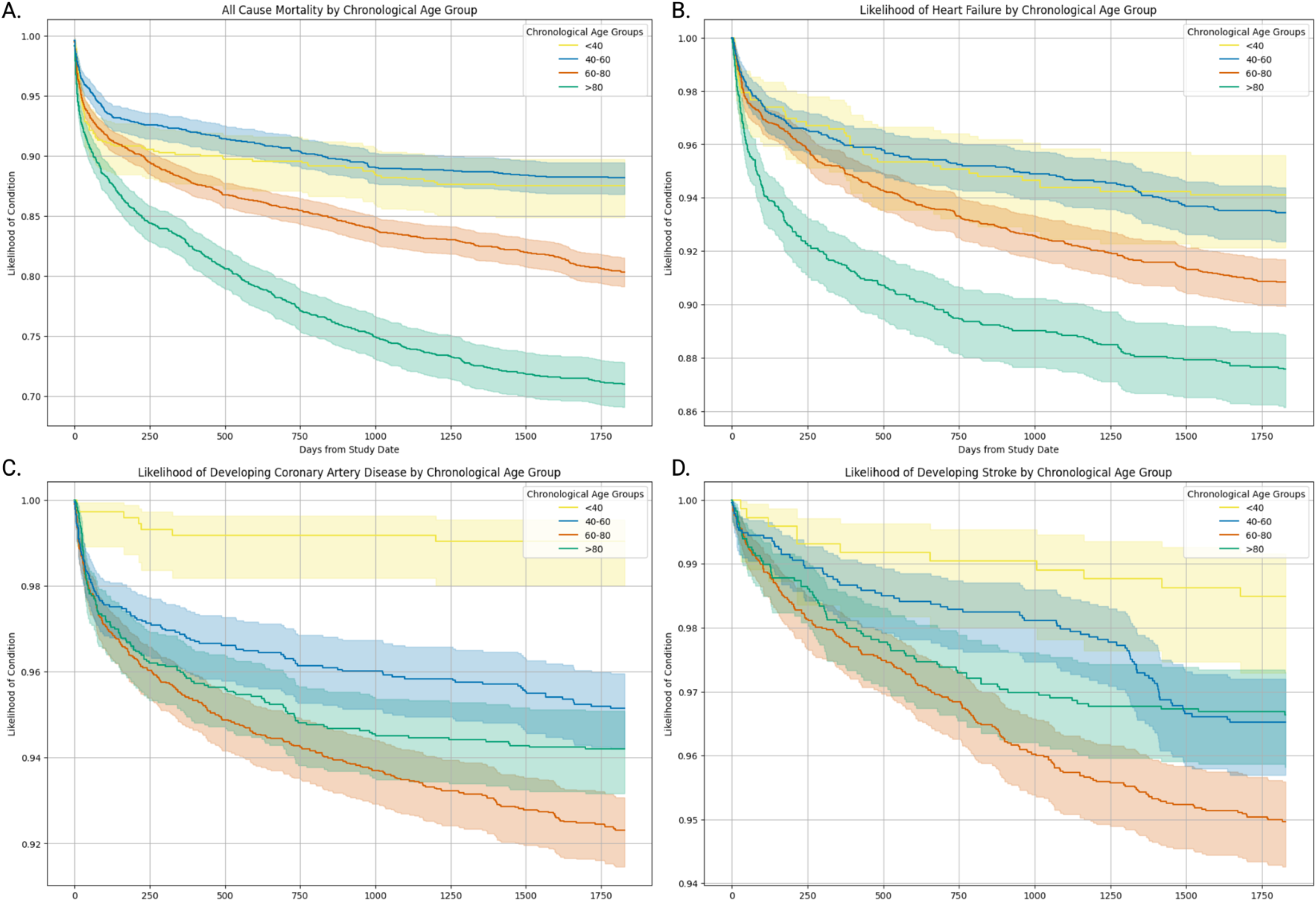
Kaplan-Meier survival curves illustrating the association between chronological age group and four clinical endpoints: (A) all-cause mortality, (B) heart failure, (C) coronary artery disease, and (D) stroke

**Supplementary Figure 2.**
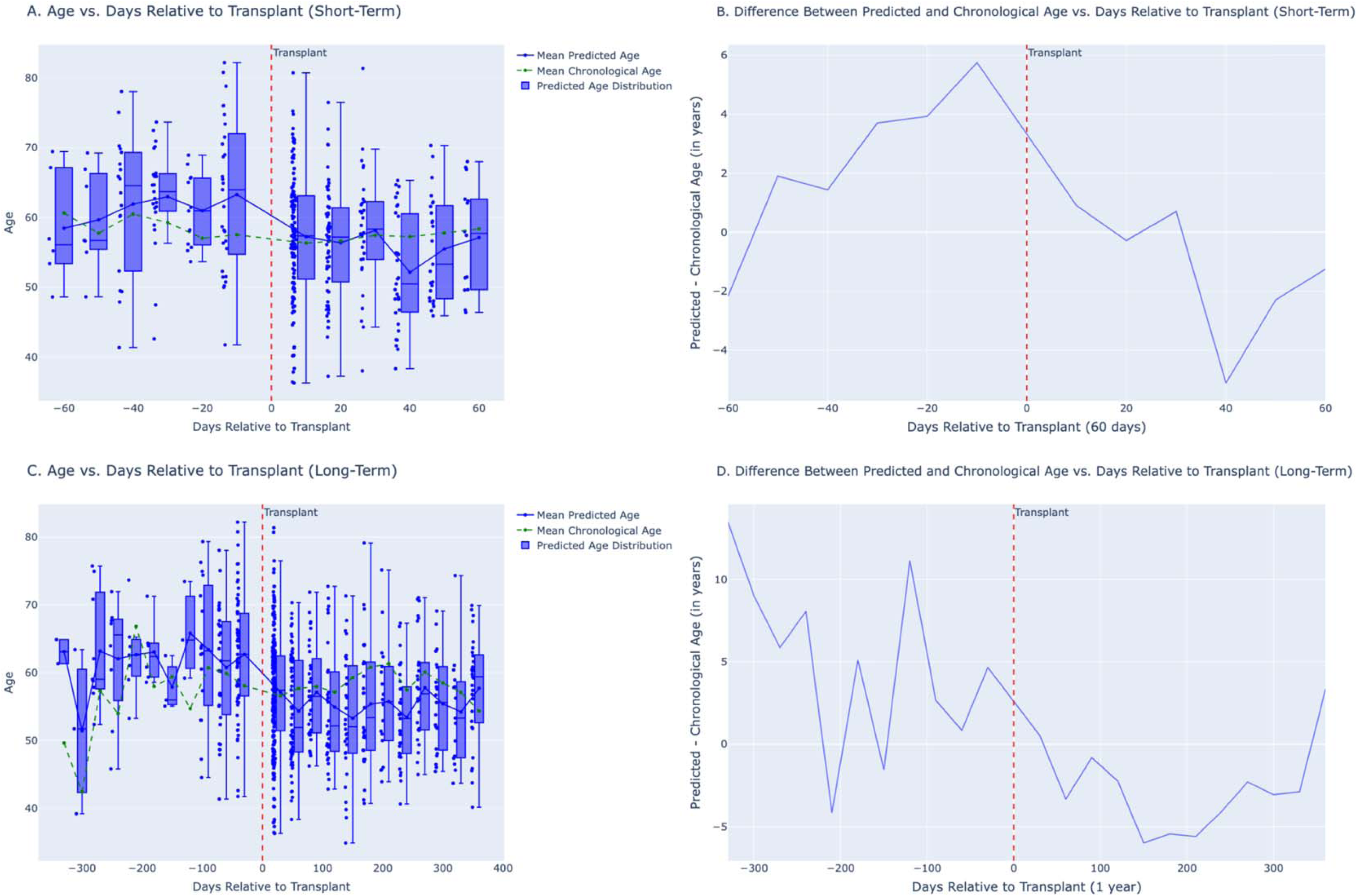
Changes in predicted age (left panels) and the difference between predicted and chronological age (right panels) over time relative to the day of transplant (red dashed line at x = 0). Days before transplant are negative on the x-axis, and days after transplant are positive. (A) and (C) show boxplots of predicted age for shorter (A) and extended (C) time windows (60 days, with 10-day interval; one year with 30-day interval). (B) and (D) show how much the predicted age differs from chronological age for shorter (B) and extended (D) time window.

**Supplementary Table 1:** Imaging characteristics grouped by predicted age group.

| Characteristic | Predicted Age Group |  |  |  |
| --- | --- | --- | --- | --- |
|  | <40 years | 40–60 years | 60–80 years | >80 years |
| Male Count (n, %) | 242 (51.49%) | 1482 (60.51%) | 2899 (61.02%) | 897 (48.30%) |
| LVEF < 41 (n, %) | 25 (5.32%) | 321 (13.11%) | 794 (16.71%) | 213 (11.47%) |
| Aortic Stenosis (n, %) | 0 (0.00%) | 23 (0.94%) | 335 (7.05%) | 434 (23.37%) |
| Mitral Regurgitation (n, %) | 59 (12.55%) | 630 (25.72%) | 1954 (41.13%) | 1133 (61.01%) |
| Tricuspid Regurgitation (n, %) | 130 (27.66%) | 869 (35.48%) | 2347 (49.40%) | 1282 (69.04%) |
| Aortic Regurgitation (n, %) | 20 (4.26%) | 204 (8.33%) | 961 (20.23%) | 712 (38.34%) |
| CABG (n, %) | 0 (0.00%) | 57 (2.33%) | 320 (6.74%) | 119 (6.41%) |
| Valve Replacement (n, %) | 1 (0.21%) | 84 (3.43%) | 442 (9.30%) | 304 (16.37%) |
| Heart Transplant (n, %) | 14 (2.98%) | 572 (23.36%) | 443 (9.32%) | 4 (0.22%) |
| Age (years) | 32.79 ± 8.36 | 52.36 ± 11.04 | 70.22 ± 10.55 | 84.98 ± 7.91 |
| Predicted Age Ensemble | 33.96 ± 3.7 | 52.45 ± 5.27 | 70.02 ± 5.64 | 84.82 ± 3.55 |
| Predicted Age PLAX | 35.68 ± 5.59 | 54.47 ± 6.86 | 69.42 ± 5.12 | 79.19 ± 3.19 |
| Predicted Age A2C | 37.42 ± 8.25 | 54.87 ± 7.91 | 69.13 ± 6.84 | 79.16 ± 5.42 |
| Predicted Age SC | 43.71 ± 7.63 | 58.49 ± 6.56 | 67.37 ± 5.27 | 74.56 ± 4.74 |
| Predicted Age A4C | 37.4 ± 6.27 | 56.51 ± 7.19 | 71.1 ± 5.74 | 80.89 ± 3.7 |
| BMI | 25.36 ± 5.53 | 28.42 ± 8.29 | 27.19 ± 6.52 | 25.57 ± 6.95 |
| Systolic BP (mmHg) | 113.97 ± 20.12 | 121.16 ± 22.7 | 122.69 ± 31.23 | 130.16 ± 47.97 |
| Diastolic BP (mmHg) | 70.7 ± 13.11 | 75.13 ± 15.44 | 69.75 ± 16.28 | 68.06 ± 15.34 |
| LVEF (%) | 60.17 ± 10.34 | 57.32 ± 14.64 | 55.87 ± 15.74 | 58.54 ± 13.03 |
| LVIDd 2D (cm) | 4.78 ± 2.59 | 4.64 ± 1.35 | 4.53 ± 0.92 | 4.28 ± 1 |
| LVIDs 2D (cm) | 3.26 ± 2.29 | 3.19 ± 1.17 | 3.18 ± 1.01 | 2.91 ± 0.75 |
| LVPWd 2D (cm) | 0.93 ± 0.38 | 1.05 ± 0.33 | 1.1 ± 0.29 | 1.16 ± 0.41 |
| LVPWs 2D (cm) | 1.3 ± 0.52 | 1.39 ± 0.53 | 1.53 ± 0.36 | 1.61 ± 0.63 |
| IVSd 2D (cm) | 0.92 ± 0.4 | 1.07 ± 0.33 | 1.14 ± 0.35 | 1.21 ± 0.29 |
| IVSs 2D (cm) | 1.42 ± 0.23 | 1.29 ± 0.34 | 1.49 ± 0.34 | 1.58 ± 0.35 |
| LA Dimension 2D (cm) | 3.24 ± 0.64 | 3.66 ± 0.81 | 3.95 ± 0.85 | 4.12 ± 0.88 |
| TR Peak Gradient (mmHg) | 19 ± 8.1 | 23.97 ± 12.08 | 27.23 ± 11.91 | 32.17 ± 12.95 |
| RA Pressure (mmHg) | 3.86 ± 2.41 | 4.83 ± 3.71 | 5.69 ± 4.26 | 5.79 ± 4.18 |
| PA Pressure (mmHg) | 22.85 ± 8.43 | 28.6 ± 12.03 | 33.2 ± 13.71 | 38.34 ± 14.46 |
| Total Count (n) | 470 | 2449 | 4751 | 1857 |

**Supplementary Table 2:** ICD codes used for exclusion criteria.

| Disease | ICD-9 Codes | ICD-10 Codes |
| --- | --- | --- |
| CABG | V45.81, V45.09, 414.01, 414.05, 996.03 | Z95.1, Z95.5, I25.10, I25.700, I25.708, I25.709, T82.211A, T82.218A, T82.218D, T82.218S |
| Valve Replacement | V15.1, V42.2, V43.3 | Z95.2, Z95.3, Z95.4, Z95.4 |
| Heart Transplant | V42.1 | Z94.1 |

